# Auditory sustained potential as a biomarker of language functioning in children with autism and its implication in clinical trials

**DOI:** 10.1101/2025.10.22.25338568

**Authors:** Vardan Arutiunian, Saeideh Davoudi, Ghizlane Gaougaou, Inga Sophia Knoth, Rae Buckser, Valérie Marcil, Sarah Lippé

**Affiliations:** Azrieli Research Center of CHU Sainte-Justine, 3175 Côte Sainte-Catherine, Montreal, QC H3T 1C5, Canada; Department of Psychology, Université de Montréal, QC H3T 1A8, Canada; Department of Nutrition, Université de Montréal, Montreal, QC H3T 1A8, Canada

## Abstract

Language impairment is the most frequently reported co-occurring condition in autism; however, its neural mechanisms are not well understood. A potential neural biomarker associated with language in autism can be a 40Hz Auditory Steady-State Response (ASSR) triggered by periodic click trains which in electroencephalogram (EEG) evokes two types of responses – *40Hz steady-state gamma response* (or ASSR) and *sustained potential* (SP). Although these responses represent low-level auditory processing, they correspond to different stages of sound perception/analysis and are essential in processing of spectrally/temporally complex sounds, including speech. However, until now there were no studies focusing on the potential of these responses to serve as objective measures in clinical trials. This open-label clinical trial evaluated the effects of the probiotic beverage supplement Bio-K+ in children with autism. Participants were assessed at three timepoints: T0 (baseline), T14 (14 weeks after treatment initiation), and T22 (8 weeks post-treatment, during the ‘wash-out’ phase), including EEG 40Hz ASSR and behavioral phenotyping. *First,* at T0 we showed a reduction of SP amplitude in children with autism compared to typically-developing (TD) controls, and this reduction was associated with lower language skills. *Second,* the amplitude of SP significantly changed during the treatment period: by T14 it became similar to that of TD children. *Finally,* these changes in the amplitude of SP were associated with improvement in language skills. Importantly, we showed that this biomarker as well as its change was related specifically to language, but not to other behavioral measures.

## INTRODUCTION

Autism Spectrum Disorder (ASD) is a neurodevelopmental condition affecting approximately 1 out of 31 children (1) and being diagnosed based on behavioral symptoms in the domains of social communication and restricted and repetitive behaviors (2). Although language impairment is not a diagnostic criterion of ASD, about 75% of autistic children have language difficulties, varying in severity, i.e., from normal language skills (∼25%) to severe language impairments with minimal or no verbal skills (3–6). However, neural mechanisms underlying these difficulties remain largely unexplored, and there is a need to identify reliable biomarkers of language impairment in ASD that can serve as objective measures in clinical trials.

It has been demonstrated that altered low-level sensory processing in the auditory cortex accounts for alterations in high-level communication skills in children with ASD, including language (7–12). One of the reliable paradigms to register this low-level sensory processing is 40Hz Auditory Steady-State Response (40Hz ASSR), where participants are presented with a click train or amplitude-modulated tones at a gamma frequency range (30–80Hz in magneto- or electroencephalography, MEG or EEG), usually at ∼ 40Hz (13–15). Previous studies have shown that such stimuli activate neuronal populations in the primary and secondary auditory cortices and elicit two distinct EEG responses: (1) an oscillatory response at the stimulation frequency (typically 40 Hz), known as the ASSR, which is commonly quantified using inter-trial phase coherence (ITPC); and (2) a sustained potential (SP), characterized by a stable negative deflection in the evoked potential during sound presentation (13,16–19). The advantage of the ASSR paradigm is that it is a passive methodology, requiring no response from participants and has a high test-retest reliability (20,21). This makes the paradigm well-suited to clinical trials, especially those which include non-verbal or minimally verbal individuals.

There are few studies on individuals with ASD using 40Hz ASSR, and the results are inconsistent. Some studies have reported a reduction of 40Hz auditory gamma response in children and adolescents with ASD, as well as in first-degree relatives of individuals with ASD (22–25) compared to typically developing (TD) controls. However, other studies have demonstrated no difference between children with and without ASD in this response (26–29). Some findings have also indicated that alterations in 40Hz ASSR can be related to language impairment in children with ASD (22,30). These inconsistencies in results can be explained by the highly heterogeneous nature of the ASD population and the variability in language skills in ASD. That is, it has been shown that the key part of the neural circuitry responsible for entrainment to exogenous auditory stimuli involves parvalbumin positive (PV+) basket cells and the pyramidal neurons of the upper layers of the auditory cortex, and the occupation of a specific channel on those PV+ cells reduced 40 Hz auditory gamma response highlighting the specific molecular mechanism contributing to the strength of 40Hz ASSR (31). Therefore, some authors suggest that the reduction of 40Hz ASSR may be associated not with the whole ASD population, but rather with a specific genetic subgroup of individuals with ASD (31,32).

Although SP is an auditory response evoked by the same periodic stimuli as the oscillatory steady-state response, it has not been widely investigated until recently. There is a very limited number of studies that have focused on this response in neurodevelopmental populations (ASD and Rett Syndrome), and they have consistently showed a reduction of the amplitude of SP (26,29,33). The authors proposed that SP is related to pitch processing as it can be triggered by any periodic spectrally complex sounds, including speech sounds (29,33,34), and, thus, the adequate functioning of SP is essential for speech perception and language processing. Therefore, a reduced SP could be a promising marker of speech/language impairments.

An important insight into the distinct cellular and system mechanisms of ASSR and SP comes from studies in animals and human neuroimaging research. Both neuronal findings in monkeys (35) and MEG studies in children and adults (18,29) have shown similar results, such that the anatomical sources of ASSR and SP are distinct: while both auditory responses are generated by Heschl’s (transverse temporal) gyrus, oscillatory ASSR at a specific stimulation frequency is generated by the primary auditory cortex (A1) whereas the localization of SP is in the anterolateral region of Heschl’s gyrus (34), which represents a so-called ‘pitch processing center’ (36,37). Evidence from electrophysiological recordings in animals suggested two distinct types of neurons in those regions with a wide representation of non-synchronized cell populations in the ‘pitch processing center’ that generates SP (35). This highlighted the differences in functional characteristics of ASSR and SP as these responses are generated not only by different areas of the auditory cortex, but also by different cell types. Given these differences, ASSR and SP reflected different stages of sound processing, where ASSR is a primary neural response at a specific frequency of stimulation and SP is a more complex response that integrates pitch information across different frequency units (35). It is important to note, however, that despite morphological and functional differences these neural responses reflect low-level auditory processing (38).

Although SP can have relevance for speech perception / language skills (26,29,33) and can be acquired from non-verbal / minimally verbal children, it has never been investigated in clinical trials. We aim to fill this gap and to explore, for the first time, SP as a neural biomarker with group discriminative potential, its relation to language skills and its changes during treatment in association with the changes in behavioral language skills. If SP is a sensitive biomarker of language skills variation in ASD, it should also be sensitive to individual variations resulting from treatment.

The present study is a 30-week, open-label, non-randomized clinical trial aiming to test the acceptability and the safety of the 3-strains probiotic beverage supplement (Bio-K+) and the feasibility of the proposed protocol in children with ASD (see [39] for the full protocol and study design). It includes multiple EEG-based measures collected at three timepoints (T0 = baseline, T14 = 14 weeks after the beginning of the treatment, T22 = 8 weeks after the end of the treatment, ‘wash-out’ stage), including ASSR paradigm. In the framework of the present study, we had several goals:

- To provide between-group comparisons (ASD vs. TD) in both 40Hz ASSR (measured with ITPC) and SP at the baseline timepoint (T0).
- To explore relationships between auditory responses (40Hz ASSR and SP) and behavioral assessments of language skills as well as other cognitive, sensory, and health measures in the ASD group at the baseline timepoint (T0).
- *Treatment effect:* to investigate the changes of 40Hz ASSR and SP after 14 weeks from the start of Bio-K+ probiotic product administration (T14) and at the post-treatment timepoint (T22) in comparison to baseline (T0) in the ASD group. Also, to reveal whether the changes in the auditory responses (40Hz ASSR and SP) were related to changes in behavioral measures in the ASD group during the treatment period (from T0 to T14).

## MATERIALS AND METHODS

### Participants

A total of 128 EEG files (64 EEGs for 40Hz ASSR and 64 EEGs for SP) for three timepoints were analyzed, resulting in 19 children with ASD (5 female, age range 4–11 years, *M* = 7.10) at three timepoints and 10 TD controls (5 female, age range 4–11 years, *M* = 7.8) at one timepoint. There were no between-group differences in age, *t*(17.9) = –0.78, *p* = 0.44, and sex, χ^2^(1) = 0.75, *p* = 0.39.

Participants were recruited via advertisements on social media or medical charts. All children in the ASD group had a medical diagnosis of ASD based on Autism Diagnosis Observation Schedule – Second Edition (40). Exclusion criteria were (1) autism in the context of genetic syndrome such as Fragile X or tuberous sclerosis complex, (2) cancer, diabetes or genetic disorder such as Down Syndrome 21 or 14; (3) immune system disorder; (4) intolerance or allergy to the probiotic beverage; (5) having taken probiotics during the previous three months; and (6) having taken antibiotics in the previous month. TD children had no history of psychiatric and/or neurodevelopmental disorders.

This study was approved by the ethics review board of the Centre Hospitalier Universitaire Sainte-Justine (#2021–3412). Informed consent was obtained from all parents involved in the study and participants gave their assent when possible.

### Bio-K+ probiotic supplement administration

Bio-K+ is a brand of commercially available and well-defined specific probiotic food that consists of a vegan pea-based, raspberry-flavored fermented drinkable product containing a minimum of 50 × 109 colony forming units (CFU) of three strains: *L. acidophilus* CL1285, *L. casei* LBC80R and *L. rhamnosus* CLR2. The product was manufactured and graciously supplied by Kerry Canada inc. (Laval, Quebec, Canada). For the duration of the supplementation period, children with ASD consumed one bottle per day (98 g) all at once or over the course of the day. Children were allowed to mix the content of the probiotic beverage bottle with any cold beverage. See more about acceptability and safety of the Bio-K+ probiotic food administration in the full protocol (39).

### Behavioral assessment

Each participant’s behavioral phenotype was assessed with standardized tools. We utilized the Autism Treatment Evaluation Checklist (41) completed by parents and used scores in the three domains: a) speech / language / communication; b) sociability; c) sensory / cognitive awareness, where higher scores indicate greater symptom severity. This checklist has been specifically developed to assess change in existing symptoms by comparing scores over time, making it suitable for clinical trial studies. Gastrointestinal (GI) symptoms were screened with the Gastrointestinal Severity Index (42), a questionnaire that measures nine components of GI distress, where higher scores indicate greater symptom severity. Sleep issues were assessed via the Children’s Sleep Habit Questionnaire (43), where a total score of 41 or more indicates that symptoms are clinically significant.

### Experimental paradigm, EEG data collection and analysis

In total, each participant was presented with 50 auditory stimuli that evoked 40Hz ASSR in a random order with the other stimuli (n = 50, at another frequency, 6Hz). Participants were exposed to click trains (1.5 ms bursts of white noise, see [44]) presented at a frequency of 40 per second (40Hz) with a random inter-stimulus interval of 1–1.5 seconds. Auditory stimuli were presented through dual speakers positioned one meter from the participant on either side. As the auditory processing task did not require active attention, participants watched a movie with the sound muted and without subtitles during the recording to maintain wakefulness and minimize head movements.

At all timepoints, EEG was acquired with a high-density 128-channel net from Electrical Geodesics, Inc. (Magstim EGI, Eugene, OR, USA) in a darkened, soundproof room. Signal was acquired using an EGI Net Amp 300 amplifier and saved on a G4 Macintosh desktop computer using NetStation EEG Version 4.5.4. Data were collected at a 1000Hz sampling rate, the vertex (Cz) was used as an online reference electrode, and impedances were maintained at 40 kΩ or below. To provide standardized EEG data processing, we used the Harvard Automated Preprocessing Pipeline for EEG (HAPPE) for artifact detection, cleaning and rejection (45) developed specifically for pediatric populations with neurodevelopmental disorders and high- artefact data. After artefact removal and correction, 6 out of 128 EEG recordings were excluded from the analysis due to severely impacted signals. Table 1 presents the EEG quality metrics for participants who were included in the analysis (with valid EEG data after pre-processing).

**Table 1.**
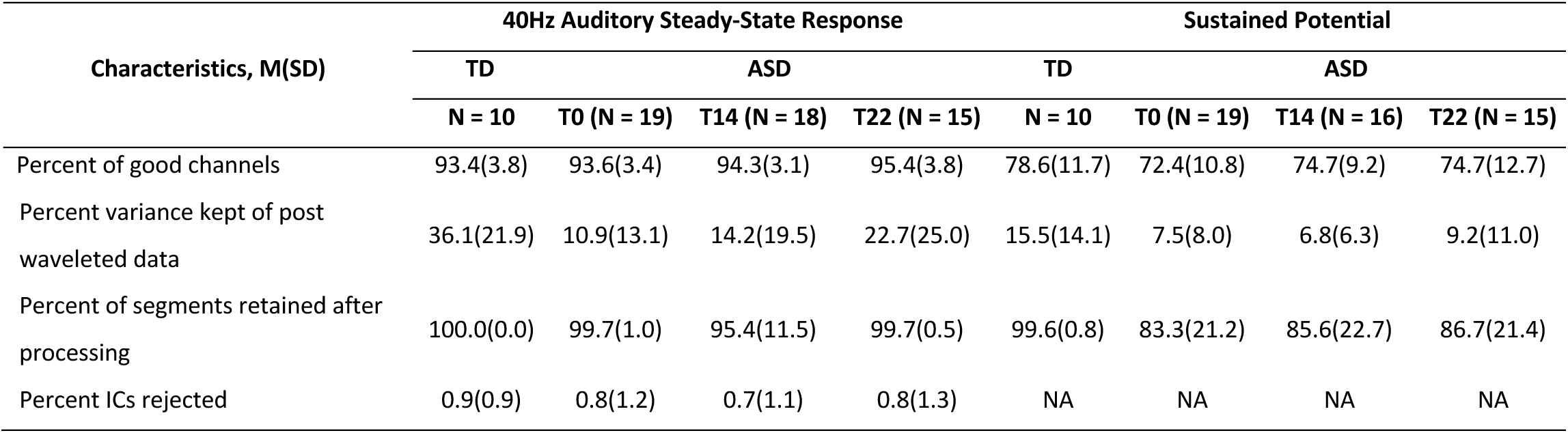
EEG quality metrics for participants who were included in the analysis (with valid EEG data after pre-processing).

Offline band-pass filters of 1–100Hz for time-frequency analysis (to calculate 40Hz ITPC) and 0.1–9Hz for evoked potential analysis (to calculate SP) were used. The cleaned EEG data were cut in 3000ms epochs (ranging from –1500 to 1500ms) for 40Hz ITPC calculation and in 1600ms epochs (ranging from –300 to 1300ms) for SP calculation; baseline correction from – 299 to 0ms was applied for both types of the analyses. To estimate 40Hz ITPC, we averaged ITPC-values in 0–1000ms time interval (stimulus presentation time) in 39–41Hz frequency range; ITPC can be values from 0.0 to 1.0, where higher values indicate higher consistency of phases across the trials (13). It has been shown that ITPC is a more reliable measure compared to the total power in the same frequency range (22). To estimate SP, we averaged the amplitude of this evoked potential in 200–600ms after stimulus onset. This time window was chosen based on the very limited number of previous studies showing that the sustained part of the evoked response starts at around 200ms after stimulus onset and the initial part of this response (but not the later) is altered in ASD (23,26,29). Both 40Hz ITPC and SP were calculated for the Fz electrode (#11 in Magstim EGI system) which captures auditory potentials on the scalp EEG (46).

### Statistical analysis

Statistical analysis was done in R (47), using *lme4* package (48); correction for multiple comparisons (false discovery rate, FDR) was applied to each set of the analyses, and *p*-values were corrected with the *p.adjust.method* in R. The data were plotted with *ggplot2* (49), and the tables for model outcomes (Tables 2–4) were created with the *sjPlot* package (50). Figures representing neural responses were created with the python data visualization library *matplotlib* (51) for time-frequency maps and with the EEGLAB (52) for evoked potentials. The structure of the models will be specified further in the Results section.

**Table 2.** Changes in the scores of the main behavioral measures during treatment (T0 = baseline / treatment’s start, T14 = 14 weeks after T0 / treatment’s end, T22 = 8 weeks after T14, post-treatment time). Significance is labeled with \**p* < 0.05, \*\**p* < 0.01, \*\*\**p* < 0.001 (significant *p*-values are FDR-corrected) and highlighted in bold.

| *Predictors* | Speech/Language/Communication |  |  |  | Sociability |  |  |  | Sensory/Cognitive awareness |  |  |  | Gastrointestinal symptoms |  |  |  | Sleep, CSHQ |  |  |  |
| --- | --- | --- | --- | --- | --- | --- | --- | --- | --- | --- | --- | --- | --- | --- | --- | --- | --- | --- | --- | --- |
|  | *Estimate* | *SE* | *t* | *p* | *Estimate* | *SE* | *t* | *p* | *Estimate* | *SE* | *t* | *p* | *Estimate* | *SE* | *t* | *p* | *Estimate* | *SE* | *t* | *p* |
| (Intercept) | 9.96 | 1.95 | 5.10 | **<0.001\*\*\*** | 11.13 | 1.45 | 7.66 | **<0.001\*\*\*** | 11.68 | 1.46 | 8.02 | **<0.001\*\*\*** | 4.61 | 0.55 | 8.42 | **<0.001\*\*\*** | 49.30 | 2.05 | 24.04 | **<0.001\*\*\*** |
| Timepoint, T14 | -2.02 | 0.80 | -2.51 | **0.019\*** | -3.83 | 1.01 | -3.78 | **0.001\*\*** | -4.17 | 1.10 | -3.80 | **0.001\*\*** | -3.16 | 0.56 | -5.59 | **<0.001\*\*\*** | -5.04 | 1.51 | -3.33 | **0.003\*\*** |
| Timepoint, T22 | -0.87 | 0.86 | -1.02 | 0.316 | 0.54 | 1.08 | 0.50 | 0.620 | -0.97 | 1.17 | -0.83 | 0.411 | -0.26 | 0.59 | -0.44 | 0.660 | -1.95 | 1.56 | -1.25 | 0.220 |
| **Random effects** |  |  |  |  |  |  |  |  |  |  |  |  |  |  |  |  |  |  |  |  |
| σ<sup>2</sup> | 4.47 |  |  |  | 7.23 |  |  |  | 8.55 |  |  |  | 2.48 |  |  |  | 16.11 |  |  |  |
| τ<sub>00 ID</sub> | 69.83 |  |  |  | 31.90 |  |  |  | 30.22 |  |  |  | 3.20 |  |  |  | 56.67 |  |  |  |
| ICC | 0.94 |  |  |  | 0.82 |  |  |  | 0.78 |  |  |  | 0.56 |  |  |  | 0.78 |  |  |  |
| N<sub>ID</sub> | 20 |  |  |  | 20 |  |  |  | 20 |  |  |  | 21 |  |  |  | 19 |  |  |  |
| Observations | 45 |  |  |  | 45 |  |  |  | 45 |  |  |  | 48 |  |  |  | 45 |  |  |  |
| Marginal R<sup>2</sup> / | 0.010 / |  |  |  | 0.091 / |  |  |  | 0.081 / |  |  |  | 0.271 / |  |  |  | 0.059 / |  |  |  |
| Conditional R<sup>2</sup> | 0.940 |  |  |  | 0.832 |  |  |  | 0.797 |  |  |  | 0.681 |  |  |  | 0.792 |  |  |  |

**Table 3.** Associations between 40Hz Inter-Trial Phase Coherence (ITPC) and behavioral measures in children with Autism Spectrum Disorder (ASD) at T0. The relation of T0-T14 change value of 40Hz ITPC and T0-T14 change scores of behavioral measures in children with ASD. Significance is labeled with \**p* < 0.05, \*\**p* < 0.01, \*\*\**p* < 0.001 (significant *p*-values are FDR-corrected) and highlighted in bold.

| *Predictors* | **40Hz Inter-Trial Phase Coherence** |  |  |  | **40Hz Inter-Trial Phase Coherence, T0-T14 change value** |  |  |  |  |
| --- | --- | --- | --- | --- | --- | --- | --- | --- | --- |
|  | *Estimate* | *SE* | *t* | *p* | *Predictors, T0-T14 change score* | *Estimate* | *SE* | *t* | *p* |
| (Intercept) | 0.11 | 0.24 | 0.48 | 0.645 | (Intercept) | -0.01 | 0.06 | -0.23 | 0.823 |
| Language | -0.02 | 0.01 | -1.85 | 0.097 | Language | 0.02 | 0.02 | 1.04 | 0.327 |
| Sociability | 0.01 | 0.01 | 0.90 | 0.392 | Sociability | 0.00 | 0.01 | 0.44 | 0.673 |
| Sensory/Cognitive | -0.00 | 0.01 | -0.30 | 0.770 | Sensory/Cognitive | -0.01 | 0.01 | -0.92 | 0.383 |
| Gastrointestinal symptoms | 0.00 | 0.02 | 0.09 | 0.930 | Gastrointestinal symptoms | -0.00 | 0.01 | -0.41 | 0.696 |
| Sleep | 0.01 | 0.01 | 1.09 | 0.305 | Sleep | -0.00 | 0.01 | -0.32 | 0.757 |
| Observations | 15 |  |  |  | Observations | 14 |  |  |  |
| R<sup>2</sup> / R<sup>2</sup> adjusted | 0.451 / |  |  |  | R<sup>2</sup> / R<sup>2</sup> adjusted | 0.149 / - |  |  |  |
|  | 0.146 |  |  |  |  | 0.382 |  |  |  |

**Table 4.** Associations between the amplitude of sustained potential (SP) and behavioral measures in children with Autism Spectrum Disorder (ASD) at T0. The relation of T0-T14 change value of the SP amplitude and T0-T14 change scores of behavioral measures in children with ASD. Significance is labeled with \**p* < 0.05, \*\**p* < 0.01, \*\*\**p* < 0.001 (significant *p*-values are FDR-corrected) and highlighted in bold.

| *Predictors* | **Sustained Potential** |  |  |  | **Sustained Potential, T0-T14 amplitude change** |  |  |  |  |
| --- | --- | --- | --- | --- | --- | --- | --- | --- | --- |
|  | *Estimate* | *SE* | *t* | *p* | *Predictors, T0-T14 change score* | *Estimate* | *SE* | *t* | *p* |
| (Intercept) | -5.69 | 6.44 | -0.88 | 0.400 | (Intercept) | 3.55 | 3.05 | 1.16 | 0.283 |
| Language | 0.72 | 0.23 | 3.10 | **0.025\*** | Language | 3.39 | 1.08 | 3.13 | **0.033\*** |
| Sociability | 0.25 | 0.21 | 1.19 | 0.266 | Sociability | -0.19 | 0.38 | -0.51 | 0.623 |
| Sensory/Cognitive | -0.41 | 0.28 | -1.47 | 0.176 | Sensory/Cognitive | -1.35 | 0.54 | -2.50 | 0.082 |
| Gastrointestinal symptoms | 0.55 | 0.52 | 1.06 | 0.316 | Gastrointestinal symptoms | 0.14 | 0.60 | 0.23 | 0.823 |
| Sleep | -0.07 | 0.14 | -0.48 | 0.641 | Sleep | -0.14 | 0.28 | -0.52 | 0.622 |
| Observations | 15 |  |  |  | Observations | 13 |  |  |  |
| R<sup>2</sup> / R<sup>2</sup> adjusted | 0.740 / |  |  |  | R<sup>2</sup> / R<sup>2</sup> adjusted | 0.618 / |  |  |  |
|  | 0.596 |  |  |  |  | 0.346 |  |  |  |

## RESULTS

### Changes in main behavioral measures during treatment

In order to investigate whether the administration of Bio-K+ probiotic food potentially improves clinical and behavioral characteristics of children with ASD, we fitted linear mixed-effects models with behavioral measures as dependent variables, timepoints as predictors (fixed effects) and participants as a random intercept. The models compared scores at T14 and T22 with the scores at T0 (reference). The results showed that after 14 weeks of intervention (by T14), there was a significant decrease in the scores for all measures, suggesting an improvement in autistic behaviors and characteristics; however, no such difference was detected at T22 (Table 2, Figure 1A).

**Figure 1.**
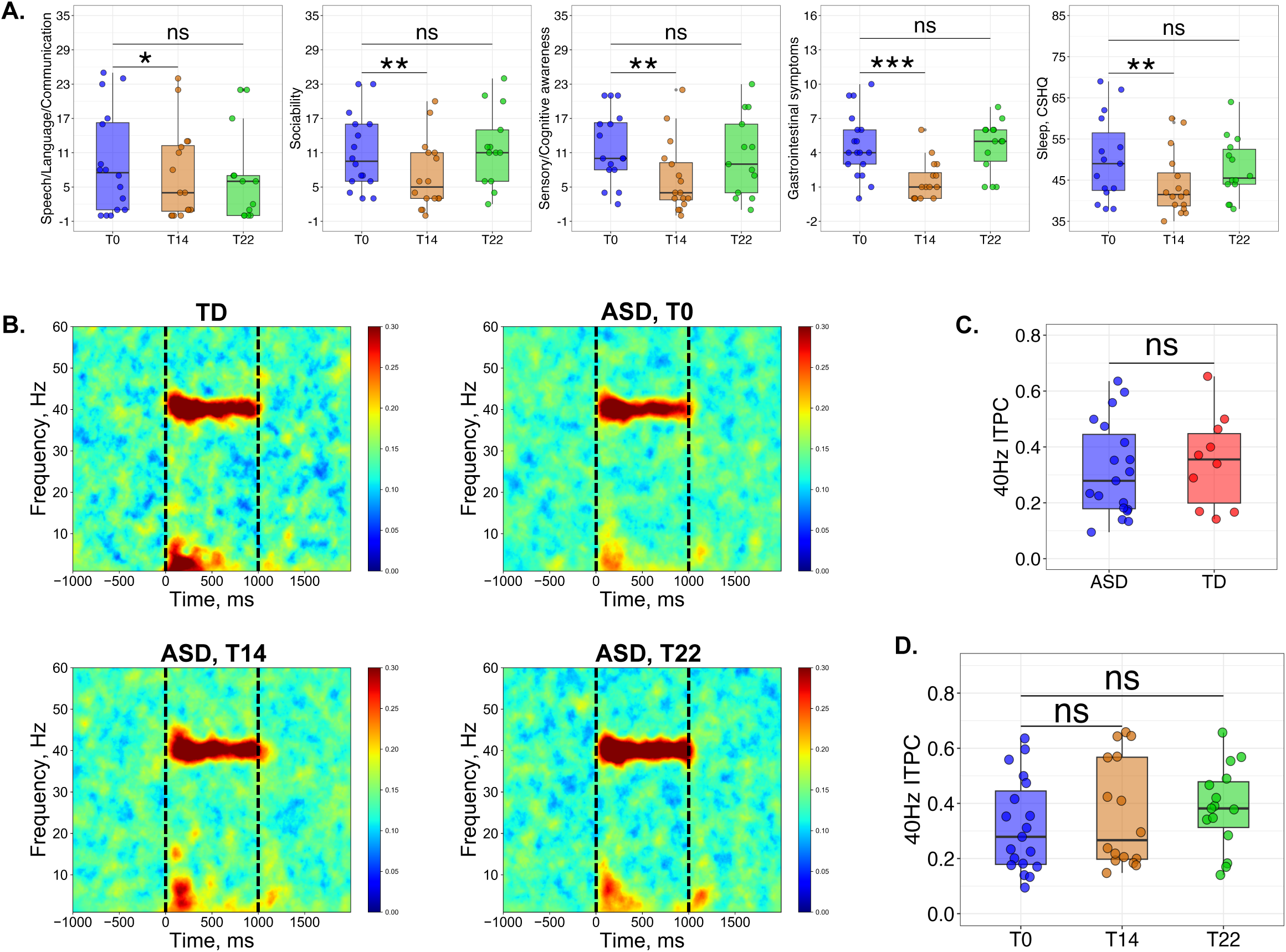
40Hz Auditory Steady-State Response (ASSR) in children with Autism Spectrum Disorder (ASD) and typically-developing (TD) controls; changes in behavioral measures during treatment in children with ASD: **A.** Differences in behavioral measures at three timepoints in children with ASD; **B.** Time-frequency maps for both groups of children at all timepoints (ASSR Inter-Trial Phase Coherence, ITPC), dashed lines represent stimulus start and end; **C.** Between-group differences in 40Hz ITPC at T0; **D.** Differences in 40Hz ITPC at three timepoints in children with ASD. Significance is labeled with \**p* < 0.05, \*\**p* < 0.01, \*\*\**p* < 0.001, ns = non-significant (significant *p*-values are FDR-corrected).

### 40Hz ASSR: group difference, relation to clinical phenotype, and changes during treatment

To provide between-group comparison in 40Hz ITPC at T0, we used Wilcoxon rank-sum test. The results did not show a significant difference, *M*_ASD_ = 0.31 (*SD* = 0.17) vs. *M*_TD_ = 0.34 (*SD* = 0.16), *W* = 95, *p* = 0.67, 95% C.I. [–0.18, 0.10], suggesting non-altered auditory gamma response in children with ASD (Figure 1B,C).

To test whether 40Hz ITPC in the ASD group at T0 was associated with behavioral measures, we fitted a liner model with the neural response as dependent variable and included five predictors (behavioral assessment of language, sociability, sensory/cognitive awareness, GI, and sleep) as main effects. The results demonstrated no significant relationships for any measures (Table 3).

To explore whether the administration of Bio-K+ probiotic food influenced 40Hz ASSR in children with ASD, we utilized a liner mixed effect model consisted of 40Hz ITPC as a dependent variable, timepoint as predictor as fixed effects and participants as a random intercept. The results did not show any changes in the auditory gamma response at any timepoint when comparing to T0: T14, Est. = 0.03, SE = 0.02, *t* = 1.68, *p* = 0.103; T22, Est. = 0.02, SE = 0.02, *t* = 1.38, *p* = 0.178 (Figure 1D).

Finally, we tested if the changes in 40Hz ITPC were related to changes in behavioral measures in children with ASD during treatment time (from T0 to T14). The results did not reveal significant relationships between variables (Table 3).

To summarize, the analysis of 40Hz ASSR – low-level auditory gamma response – showed neither a between-group (ASD vs. TD) difference, nor a relationship to clinical phenotype and changes during the administration of Bio-K+ probiotic food in children with ASD.

### Sustained potential (SP): group difference, relation to clinical phenotype, and changes during treatment

The amplitude of SP was significantly reduced in children with ASD in comparison to TD controls, *M*_ASD_ = –2.79 (*SD* = 6.02) vs. *M*_TD_ = –6.29 (*SD* = 3.26), *W* = 140, *p* = 0.03, 95% C.I. [0.24, 5.93], although the shape of the wave was similar to that in TD children, with the clear presence of the obligatory transient auditory components (Figure 2A–C).

**Figure 2.**
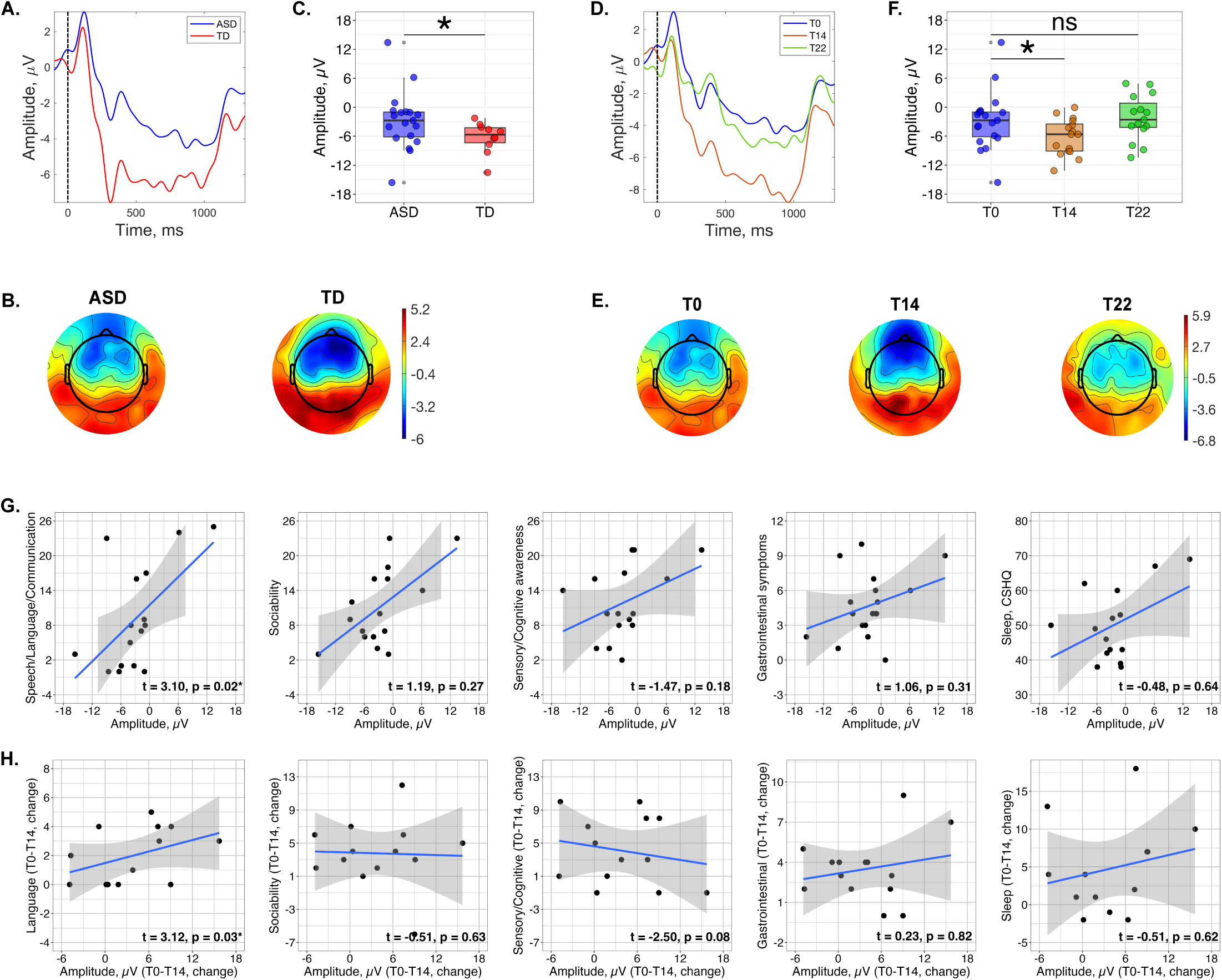
Auditory sustained potential (SP) in children with Autism Spectrum Disorder (ASD) and typically-developing (TD) controls: **A.** Timecourses of SP in both groups of children at T0 (dashed line represents the stimulus onset); **B.** The topographic distributions show the mean amplitude of SP in both groups of children in the 200–600 ms time window at T0; **C.** Between-group differences in the amplitude of SP at T0; **D.** Timecourses of SP at three timepoints in children with ASD (dashed line represents the stimulus onset); **E.** The topographic distributions show the mean amplitude of SP at three timepoints in children with ASD in the 200–600 ms time window; **F.** Differences in the amplitude of SP at three timepoints in children with ASD; **G.** Relationships between SP and behavioral measures in children with ASD at T0; **H.** Relationships between changes in SP and behavioral measures [T0-T14] in children with ASD. Significance is labeled with \**p* < 0.05, \*\**p* < 0.01, \*\*\**p* < 0.001, ns = non-significant (significant *p*-values are FDR-corrected).

To explore whether the reduction of the SP amplitude had a clinical relevance in children with ASD, we provided brain-behavior modeling at T0 and fitted a liner model with the neural response as dependent variable and included five predictors (behavioral assessment of language, sociability, sensory/cognitive awareness, GI, and sleep) as main effects. The results showed a statistically significant relationship between the amplitude of SP and language skills, Est. = 0.72, SE = 0.23, *t* = 3.10, *p* = 0.025 (Figure 2G, Table 4), indicating that the larger amplitude reduction was associated with lower language skills. Other predictors were not significant (see Table 4).

The next step of the analysis aimed to investigate a possible impact of the Bio-K+ probiotic product administration on the amplitude of SP in children with ASD. We used a linear mixed-effects model with SP as a dependent variable, timepoint as predictor as fixed effects and participants as a random intercept. The results demonstrated a significant increase in the amplitude of the auditory response by T14 (i.e., after 14 weeks of intervention, end of the treatment period) in comparison to T0: Est. = –3.51, SE = 1.34, *t* = –2.61, *p* = 0.027 (Figure 2D– F). No such change was detected for T22 when compared to T0, Est. = 0.75, SE = 1.37, *t* = 0.55, *p* = 0.586 (Figure 2D–F). This suggests that the amplitude of SP became significantly more negative by T14, thus, moving toward a more ‘typical’ shape (from T0 to T14, that is, during the treatment period). However, by T22 (after treatment period) it turned to a shape similar to that observed at T0.

A follow-up exploratory analysis compared the amplitudes of SP at T14 and T22 with the SP amplitude of TD children. At T14, we did not find a significant difference between the groups, *M*_ASD_ = –6.21 (*SD* = 3.73) vs. *M*_TD_ = –6.29 (*SD* = 3.26), *W* = 79, *p* = 0.98, 95% C.I. [–3.34, 3.39]. However, at T22, the ASD group similar as at T0 had a reduced amplitude of SP, *M*_ASD_ = –2.21 (*SD* = 4.68) vs. *M*_TD_ = –6.29 (*SD* = 3.26), *W* = 116, *p* = 0.02, 95% C.I. [0.58, 8.04].

Therefore, indeed, after 14 weeks of intervention (by the end of the treatment period) the amplitude of SP in children with ASD became similar to that of TD controls.

Finally, we tested whether the changes in the amplitude of SP were related to changes in behavioral measures in the ASD group during the time of Bio-K+ probiotic product administration (from T0 to T14). The output of the model is presented in Table 4. The results showed a significant relationship between the changes in the amplitude of SP and changes in language scores, such that the larger change in the SP toward more negative amplitude (and, thus, toward more ‘typical’ shape) was associated with the improvement in language scores, Est. = 3.39, SE = 1.08, *t* = 3.13, *p* = 0.033 (Figure 2H). Other relationships were not significant.

In summary, the analysis of the SP amplitude revealed, first, its reduction in children with ASD when compared to TD controls, and this reduction was associated with lower language skills. Second, the amplitude of SP significantly changed during the treatment period: by T14 it became similar to that of TD children. Finally, these changes in the amplitude of SP were associated with changes in language skills toward improvement. Importantly, we demonstrated that this biomarker as well as its change was specifically related to language, but not to other behavioral measures.

## DISCUSSION

The present study aimed to investigate whether 40Hz ASSR and SP could serve as biomarkers of language impairment in children with ASD and could be utilized as objective measures of the changes in language skills during treatment in clinical trials. Overall, we revealed that SP is a promising biomarker associated with language skills in ASD, that SP changes with treatment (Bio-K+ probiotic food administration), and its changes are related to changes in behavioral language skills.

Between-group comparisons (ASD vs. TD) demonstrated no difference in 40Hz steady-state gamma response at T0 which corresponds to (22–25), but not to (26–29). These inconsistencies in the previous findings can be explained by the highly heterogeneous nature of the ASD population and potential molecular mechanisms that could be altered in specific subgroups of autistic individuals (31). For example, animal studies have shown that the occupancy of *N*-methyl-D-aspartate (NMDA) receptors on PV+ interneurons reduced 40Hz auditory gamma response and, thus, alterations in this neural response may be associated with a specific subgroup within the autistic cohort but not with the whole ASD population (31,32). By contrast, we revealed a difference between groups in SP with the evidence of reduced amplitude in children with ASD. Our findings confirmed the previous studies that showed a lower amplitude of SP in children with ASD (26,29) and Rett Syndrome (33).

Specific alterations in SP, but not 40Hz ASSR, can shed light into particular mechanisms of auditory processing that are impaired in ASD. Electrophysiological recordings in animals and MEG studies in children and adults consistently demonstrated that SP and 40Hz ASSR are generated by different cortical areas inside the Heschl’s gyrus: while both responses reflect low-level auditory processing (38), 40Hz ASSR is generated by the primary auditory cortex (‘core auditory area’) whereas neural generators of SP is in anterolateral region of Heschl’s gyrus which represents so-called ‘pitch processing center’ (18,29,34–37). Given these differences, ASSR reflects a primary neural response to specific frequency of stimulation (= 40Hz) / ‘pure’ tone processing based on tonotopic organization of A1, whereas SP is a more complex response that integrates pitch information across different frequency units (35) and can be more essential for speech perception and processing in comparison to ASSR.

Indeed, our results showed a relationship between the amplitude of SP (but not 40Hz ASSR) and language skills in children with ASD, such that a larger reduction of the amplitude was associated with the lower language skills. A number of studies in humans using implanted electrodes and intracranial EEG with direct electrophysiological recordings in different parts of Heschl’s gyrus as well as lesion-symptom mapping studies have revealed functional differences between posteromedial (a neural generator of ASSR) vs. anterolateral (a neural generator of SP) regions of Heschl’s gyrus in relation to speech and nonspeech sound processing (53–60). For example, during naturalistic speech perception and intracranial EEG recordings across Heschl’s gyrus it has been shown that phonemic encoding increases toward anterolateral region of Heschl’s gyrus as well as this region demonstrated the highest sensitivity / preferentially responded to speech over non-speech sounds (57). It has been revealed that the posteromedial region of Heschl’s gyrus which represents the ‘core area’ / A1 is involved in the processing of basic acoustic features of sounds (including speech sounds) by decomposing them into pure tones, whereas anterolateral region of Heschl’s gyrus is involved in more complex integrative (but still low-level) sound processing (58). The observed relationship between the amplitude of SP and language skills in children with ASD was domain specific, as no associations were found between this neural response and other behavioral measures, confirming an essential role of SP in language.

Importantly, our results revealed the influence of the probiotic treatment on SP as well as on the changes between the amplitude of SP in relation to language skills in children with ASD. First, we showed that the amplitude of SP in children with ASD became similar to those of TD children with no between-group differences by T14, that is, by the end of the treatment / after 14 weeks of the start of Bio-K+ probiotic supplementation. However, at T22 (post-treatment ‘wash- out’ stage, 8 weeks after the end of the treatment), SP amplitude in the ASD group changed back to what it was at T0 with a reduced amplitude in comparison to TD children. Second, our findings demonstrated that the changes in the amplitude of SP were associated with changes of language skills by the end of the treatment (from T0 to T14). Specifically, a larger change in the SP toward a more negative amplitude (and, thus, toward more ‘typical’ shape) was related to improvements in language scores. It is important to highlight that, although all behavioral measures (speech / language / communication; sociability; sensory / cognitive awareness; GI symptoms; sleep issues) changed during treatment and improved by T14, only the changes in language skills were related to changes of the amplitude of SP, pointing to a domain-specific relationship. Our open-label pilot study aligns with previous studies demonstrating significant effects of probiotics on autistic symptomatology and co-occurring conditions (61–66). Probiotic intake has been suggested to restore microbial homeostasis and decrease neurobehavioral, GI and sleep issues in individuals with ASD (61), but this remains controversial as there is limited evidence to support such impact. It is known that changes in the gut microbiome can affect neural activity through the microbiota-gut-brain axis as the neuroactive gut metabolites can modulate brain activity directly, via the systemic circulation, or via vagal and spinal afferents (67,68). Therefore, our results indicated a promising effect of Bio-K+ probiotic with *L. acidophilus* CL1285, *L. casei* LBC80R and *L. rhamnosus* CLR2 on specific neural response in the auditory cortex associated with language skills in ASD.

In conclusion, this open-label clinical trial is the first to demonstrate of the relevance of SP as a neural biomarker of language impairment in children with ASD. The amplitude of this neural response was reduced in children with ASD and this reduction was related to language skills; the response changed toward ‘typical shape’ during the treatment and returned back to ‘atypical shape’ at the ‘wash-out’ stage, 8 weeks after the end of the treatment. Notably, the changes in the amplitude of SP toward more negative amplitude (i.e., more ‘typical’ shape) during treatment were related to improvement in language skills as reported by parents.

Therefore, our results suggest an EEG-based objective measure of language abilities in children with ASD that can be used in future clinical trials as it is sensitive to the treatment effects and can serve as a biomarker of the changes in language skills. Moreover, it can be administered in non-verbal or minimally verbal individuals with profound autism, a group with the highest intervention needs. These promising results, although in a relatively small sample size, highlighted the importance of using this biomarker in larger, randomized, double-blind, placebo-controlled clinical trial studies. Future studies should also explore its sensitivity to other pharmacological interventions that are currently under investigation in ASD, such as, for example, *arbaclofen* (69), *leucovorin calcium* (70), or *bumetanide* (71). In addition, future studies would benefit from testing this biomarker in other clinical populations to reveal whether SP is a universal neural marker of language functioning or is specific to language impairment in children with ASD. Finally, while we used parent reports for behavioral measures, future research needs to use more precise and direct assessments of language abilities.

## Data Availability

All data produced in the present study are available upon reasonable request to the authors

## Availability of data and materials

The datasets of the current study are available upon reasonable request.

## Acknowledgements

We gratefully acknowledge the participants and their families, the research nurses at CHU Sainte-Justine, and all contributors to this work. We would like to thank the DSMB members: Isabelle Soulières (Université du Québec à Montréal), Manuella Santos (Centre hospitalier de l’Université de Montréal), André Marette (Faculty of Medicine, Université Laval).

## Authors’ contribution

*Conceptualization:* VA, GG, ISK, VM, SL; *Methodology:* VA, GG, ISK, VM, SL; *Data curation:* VA, SD, ISK, RB; *Formal Analysis:* VA, SD; *Investigation:* GG, ISK; *Resources:* VM, SL; *Writing – Original Draft:* VA; *Writing – Review and Editing:* VA, SD, GG, ISK, RB, VM, SL; *Project Administration:* GG, ISK, VM, SL; *Visualization:* VA, SD; *Supervision:* VM, SL; *Funding Acquisition:* GG, VM, SL. All authors have read and agreed to the published version of the manuscript.

